# Assessment of factors associated with reproductive health seeking behaviours among adolescent mothers in Dodoma region

**DOI:** 10.1101/2024.05.10.24307202

**Authors:** Eliwedi Mbwambo, Nyasiro Gibore

**Author notes:** Corresponding author: (ETM).

## Abstract

**Background:** Adolescent pregnancy poses a significant public health concern due to its association with high levels of maternal morbidity and mortality, particularly in sub-Saharan countries. For early detection and management of obstetric danger signs, appropriate health seeking behaviour on reproductive health among adolescent mother is highly recommended so as to decrease maternal mortality and morbidity related to pregnancy. This study aims to assess the factors influencing reproductive health-seeking behaviours among adolescent pregnant mothers in Dodoma region.

**Methods:** A community cross-sectional analytical study will be conducted, involving multistage sampling to recruit 604 adolescent pregnant mothers. Face-to-face interviews using structured questionnaires will be utilized to assess predisposing, enabling, and need factors, as well as knowledge and attitudes regarding obstetric danger signs. Data analysis will be performed using SPSS software version 25. Descriptive statistics will be used to analyse background information, while inferential statistics, including chi-square and logistic regression, will determine associations between variables.

**Results:** This study will display the trend in utilization of reproductive health as well as predisposing, need and enabling factor associated with health seeking behaviour. A statistically significant variable in the final model will be identified with a 95% confidence interval and a p-value of < 0.05.

**Conclusions:** This study will provide a clear picture of the level of knowledge, attitude, and health-seeking behaviour which is crucial for preventing delay number one of seeking health care immediately after experiencing complications also gives a picture of the role of healthcare workers in promoting health among vulnerable populations.

## Introduction

Given that teenage pregnancy is recognized to carry hazards for both the mother and the newborn, it is a serious public health concern and a delicate subject. The second most common cause of death and the fourth major cause of life years with a handicap among adolescent women aged 15 to 19 worldwide are complications during pregnancy and childbirth(WHO, 2019). Globally about 13.3 million birth which is equal to 10% of all birth were born to adolescent mothers under the age of 20, half of them were from sub–Saharan Africa(*World Population Prospects 2022*, 2022).

Tanzania demographic health surveys revealed the decline in adolescent pregnancy of women age 15-19 from 27% in 2015 to 22% in 2020, where rural area account for 25% compared to urban area account for 16%. Education attains among women age 15-19 become significant in reducing adolescent pregnancy as those with no education 53% have ever had birth compared to 9% of those with secondary education or higher(Tanzania Demographic and Health Survey and Malaria Indicator Survey 2022 Final Report., 2022).

Globally maternal mortality rate decreased by 34% from 339 deaths to 223 deaths per 100,000 live births from 2000 to 2020 respectively and to attain the global reduction of maternal death by year 2030 below 70% as stated by Sustainable Development Goals annual rate reduction of 11.6% is required. (World Health Organisation, 2023). In Tanzania maternal mortality ratio (MMR) has also declined from 556/100,000 in 2015/2016 to 104 in 2022(Tanzania Demographic and Health Survey and Malaria Indicator Survey 2022 Final Report., 2022).

About health consequences, pregnancy during adolescent encounters higher risks of puerperal endometritis systemic infections, and eclampsia, compared to women aged 20 to 24 years(Ganchimeg et al., 2014). Additionally, adolescent girls aged 15–19 years account for almost 3.9 million unsafe abortions that occur each year, this also contributes to maternal mortality, morbidity, and lasting health problems(WHO, 2019). On the other hand, early childbearing poses risks for newborns as babies born to mothers under 20 years of age face higher risks of preterm delivery, low birth weight, and severe neonatal conditions(Moshi & Tilisho, 2023).

The problem of maternal mortality can be approached, minimized, and prevented using a delay model that includes delays in deciding to seek life-saving care, delays in reaching a health care facility, and delays in receiving the needed services upon reaching the facility. A study that was conducted in Tanzania Dodoma at Dodoma referral hospital showed that among 35 deaths that occurred in 2018, 7 deaths contributed to delayed decisions at the family level(Nasser et al., 2020). The first delay can be significantly addressed if women and their families recognize obstetric danger signs and take appropriate health-seeking action immediately (Khanom et al., 2019; Nurgi et al., 2017; Wassihun et al., 2020).

Complications related to pregnancy and childbirth are difficult to predict but can be prevented, with well-timed decisions and access to emergency maternal health care. Major obstetric complications that lead to death are eclampsia and sepsis(Nassoro et al., 2020), which can be manifested through signs such as swelling, severe headache, and visual disturbance, so the ability of the adolescent pregnant mothers to observe these danger signs and take appropriate health-seeking action immediately will result into positive pregnant outcome to both mother and newborn(Hackett et al., 2019; Kassa et al., 2019; Mph et al., 2014).

Adolescent often do not seek health care services because of their poor attitude and lack of knowledge as well as unfriendly treatment from health care provider and the community at large(Atuyambe et al., 2008; Mwilike et al., 2018). Social stigma and violence from family member and partners are also causes of the poor health seeking behaviour among adolescent mothers. During pregnancy health care seeking behaviour is highly associated with knowledge regarding obstetric danger signs, the study done in Ethiopia shown that those women who had experienced obstetric danger signs had appropriate health seeking behaviour (Yosef & Tesfaye, 2021).

In a study that was done in Tanzania shown that adolescent mother had inadequate knowledge on sexual and reproductive health as they do not attend antenatal and reproductive health program(Mpimbi et al., 2022). This in turn affect their knowledge regarding obstetric danger signs as the study done in Thailand reveal that adolescent poor have poor knowledge regarding obstetric danger signs and this was the similar to the mixed study done in Tanzania where adolescent pregnant mother had poor knowledge(Bwalya et al., 2018; Kayemba et al., 2023; Parvin & Rana, 2022).

Despite the decline in the maternal mortality ratio the target of UN Sustainable Development Goal (SDG) 3.1 to reduce the global maternal mortality ratio to less than 70 per 100,000 live births has not yet been reached, this means that still several women die from pregnancy, childbirth, and postnatal related complications, this includes an adolescent mother who legged behind in seeking health care due to lack of support, financial hardship, but also unfriendly treatment from health care provider.

Lack of reproductive health knowledge among young women, particularly adolescent mothers, compromises their ability to seek timely and appropriate medical attention due to delayed recognition of dangerous signs during pregnancy, delivery and postpartum. As a result, they are unable to get to medical facilities in time to avoid severe pregnancy complications that could have a negative impact on the mother, the unborn child, or the pregnancy itself (Habte & Dessu, 2023; Health et al., 2020; Mwilike, Nalwadda, et al., 2018; Mwilike, Shimoda, Oka, Leshabari, Shimpuku, et al., 2018).

Knowing symptoms associated with antenatal risks among pregnant women may result in seeking care earlier or self-advocating for more immediate treatment in health facilities (Article et al., 2021; Mwilike, Nalwadda, et al., 2018; Mwilike, Shimoda, Oka, Leshabari, Shimpuku, et al., 2018).Additionally, a mother’s attitude toward these warning signs influences her behaviour to seek medical attention, which is essential for safe motherhood. As a result, maternal deaths can be prevented if women who have problems can recognize these indicators and seek the necessary emergency obstetric treatment.

Although health seeking behaviours which is influenced by knowledge and attitudes about obstetric danger signs have a tremendous potential to reduce maternal and infant fatalities and morbidities, as well as to improve mothers’ actions toward seeking medical attention, which is essential for safe motherhood (Ali et al., 2020), its status is not well known among this vulnerable population of adolescent pregnant mother especially in Dodoma region which contribute a significant number of adolescent pregnancies in the country

This study aims to close this knowledge gap of current level of knowledge, attitudes toward danger signs, and their influence on behaviours related to seeking health care among pregnant adolescent mothers in the community so as to contribute in modification health policy and program on adolescent mother and hence reduce maternal morbidity and mortality rate in the county

**Theoretical model:** The Anderson Behavioral Model.

The Anderson Behavioral Model for health services utilization displays a theoretical structure to understand access to and utilization of health services and put into consideration the factors that impact one’s decision to either utilize or not utilize available health services (Alkhawaldeh et al, 2023; Travers et al., 2020). This theory takes into consideration three factors which are predisposing factors, enabling factors, and need factors as important factors that influence one’s health-seeking behaviours. Model allow the integration of intermediate variable that are knowledge and attitude towards obstetric danger signs as they have influence on reproductive health seeking behaviours among adolescent mothers.

Predisposing factor that put into accounts are demographic and social factors such as education, area of residence, access to health services, marital status, and occupation. As well as culture belief and norms.

Enabling factors include factors required to facilitate the utilization of the available health services among adolescent pregnant mothers, that are family support, access to health insurance, availability of health facility, distance from health facility, ANC visit, availability of community health workers, partner support, as well as source of income.

Need factors this include parity, gravity previous abortion, experience of danger signs and gestational age.

**Figure 1:**
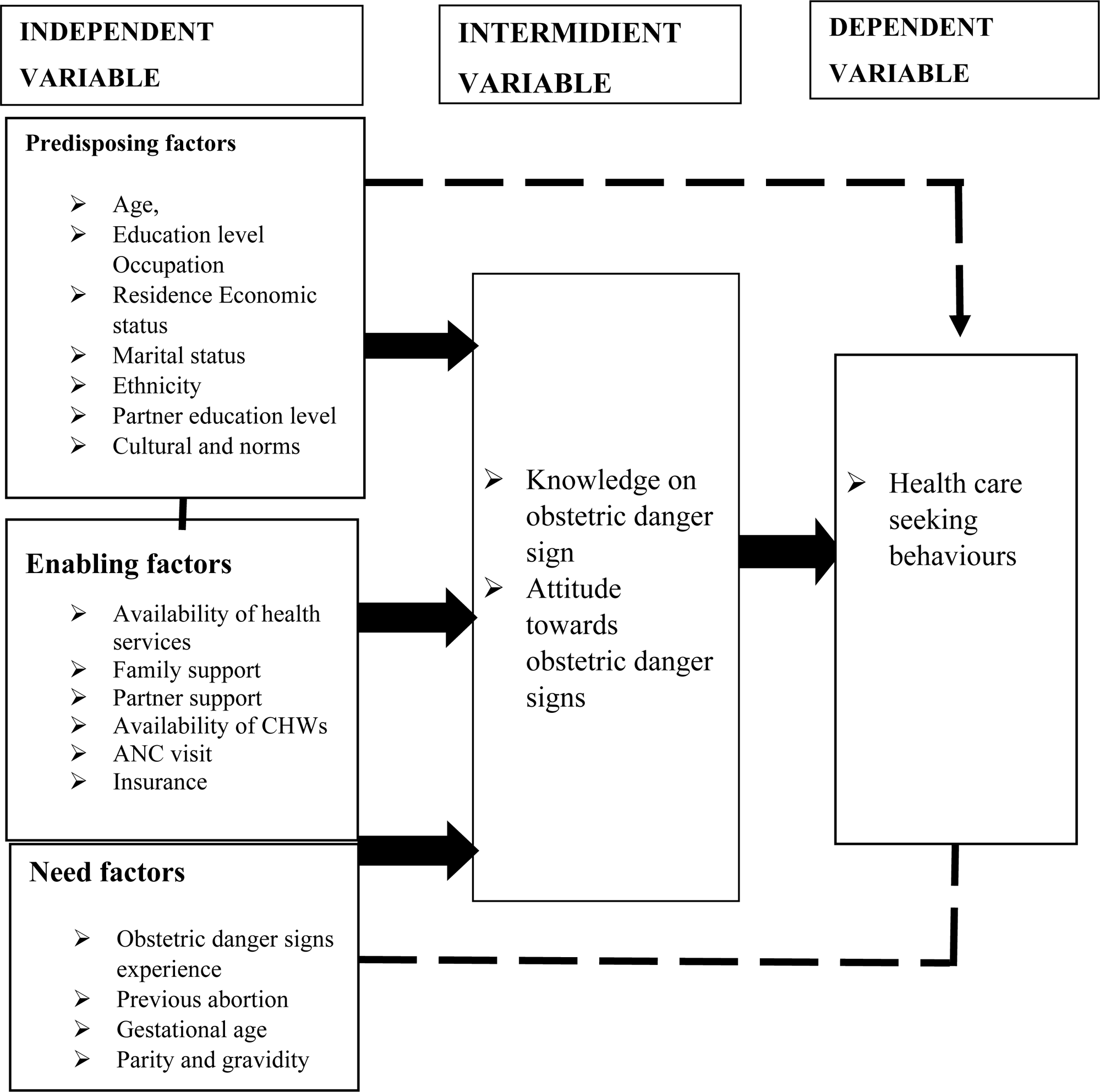
Conceptual Framework on the knowledge attitude on obstetric danger signs and their influence on health seeking behaviour.

## Method and material

### Study area

The Dodoma Region will serve as the study area. This study will be carried out in three districts—Kondoa Tc, Bahi, and Chemba—of the seven districts that make up the Dodoma region. study will be conducted from 9^th^ April to 25 May 2024.

### Study design

This study will adapt a community analytical cross-sectional design that will allows participants to be assessed only once at a point in time during the data collection period.

### Study Population

The study population will consist of adolescent pregnant mothers aged 15-19 who live in the three chosen district who will meet inclusion criteria such as, being pregnant and recent delivered mothers (42 days), signed informed consent to participate in the study for those who are above 18 years and those whose parents/caregivers signed the consent form and assent for voluntary participation in the study.

### Sample size determination

The sample size for this study will be calculated using the Kish Leslie’s formula(Rutherford, 1966), which state that N= (z^2^× p(1-p) ×d)/e^2^ this lead to sample size of 576 plus an addition of 5% for possible non-responses and error in data entry and recording this make 604.

### Sampling procedure

A three multistage sampling procedure will be used to obtain a required sample size first selection of districts second selections of wards and last village through lottery methods. Through the use of community health worker and village leaders those household with adolescent pregnant mother will be identified and those who meet criteria will be chosen convenient to serve as study sample. 50% that is 129 of total village will be included in the study whereby from each cluster (village) expected number of adolescent mothers to be included in study is five (5) this will make total of 604 a required sample size.

### Measurements of variables

#### Intermediate variable

Knowledge of obstetric danger signs will include 12 questions about danger signs occurring during pregnancy, labour, and the postpartum period. The score of knowledge variable will be in two sides good knowledge and poor knowledge. Adolescent pregnant mothers who will be able to mention 5 danger signs and 1 from each phase will be considered to have a good knowledge and those who will fail to mention 5 danger signs and 1 from each phase will be considered to have poor knowledge (Bogale & Markos, 2015; Regasa et al., 2020).

Attitude toward obstetric danger signs will include 10 questions, and it will base on a five-point Likert scale (strongly agree, agree, uncertain, disagree, and strongly disagree). Then the score will be as 5, 4, 3, 2, and 1 respectively. Negative statements will be scored in the opposite direction. Score of all eight items will be summed make 50 as highest score and 10 as a lowest score. The mean score will be calculated based on the previous studies and those who will score above the mean value will be considered to have positive attitude and those who will score below the mean score will be considered to have negative attitude.

#### Dependent variable

Health care seeking behaviours will be assessed by asking adolescent mothers at “what gestational age did they initiate ANC visit” this will include below 12weeks and above 12 weeks. Also “the actions they would take after recognizing a danger sign during pregnancy”. This will include the following behaviour response following consulting relative, her partner or a friend, self-medications, consults a traditional practitioner, doing nothing and visiting the health facility. ANC visit before 12 weeks and visit the health facility after recognition of obstetric danger signs will be considered the appropriate health seeking behaviour. The appropriate action to take will be to visit a health facility for early and prompt care and management. Other behaviours will be considered as inappropriate health seeking behaviours(Mwilike, Nalwadda, et al., 2018).

#### Independent variable

Socioeconomic and demographic characteristics, Age will be measured by one item numerical scale, and seven items in nominal scale marital status, educational level, residence, economic status, partner education level, characteristic of nearby facility and distance from nearby facility, insurance status will be measured by binary scale. Obstetric factors and antenatal factors including gestational age and number of antenatal visits will be measured by ordinal scale while history of previous abortion and previous exposure to obstetric danger signs will be measured by binary scale. Parity and gravidity will be measured by ordinal scale.

### Data collection method

Face to face interviewee will be employed for data collections. The tool will be constructed in English language and translated into Kiswahili. Four research assistance will be recruited from St John’s University, 4year students of B.Sc. Nursing then a training session of two day will be conducted. Research assistant will be taught on the purpose of the study, objectives, sampling procedure, data collection tool and method as well as informed consent.

### Data collection instrument

Structured questionnaires with closed-ended will be used for the data collection on knowledge, attitude, and health care-seeking behaviours. The survey instruments created by the JHPIEGO Maternal Neonatal Health Program will serve as the model for this tool. The following five sections will comprise it(JHPIEGO/Maternal and Neonatal Health Program, 2004). A questionnaire tool will include the following sections, section A: demographic characteristics, section B: antenatal and obstetric histories, section C: Knowledge regarding obstetric danger signs, section D: Attitude towards obstetric danger signs, section E: Health-seeking behaviours.

### Data analysis plan

The Data will be analysed using SPSS version 25. The data will be cleaned and checked for completeness before being subjected to models of analysis. Descriptive statistic will be used to analyse demographic data and frequence distribution of various items. Inferential statistic with be employed to report relationships between predisposing, enabling and need factors with health seeking behaviour. Also, to find the association between knowledge, attitude towards obstetric danger signs and health seeking behaviour inferential statistic will be used.

### Ethics statement

Ethical clearance obtained from the Ethical Review Committee of the University of Dodoma given **No: MA.84/261/02/A/72/79**, 03/04/2024. Also, permission to conduct the study obtained from relevant authorities which are the President Office Regional and Local Government Authorities (PO-RALAG), Ethical clearance later with a reference later **NO: AB 307/323/01** 08/04/2024. Regional Administrative Secretary (RAS) office issued ethical clearance with a later **NO: HA 107/249/012/109** to conduct this study in Dodoma region.

Research ethics will be put into consideration throughout the study this include, informed consent will be obtained from parents for those adolescents who still lives with their parents and are below 18 years old, assent will be provided to this group so as ensure their willingness to be part of the study. Adolescent girls starting at age 18 will be asked to provide informed consent after being fully told about the study’s purpose using language appropriate for their age.

Confidentiality will be ensured to all participants with the consideration of the potential risk of information about their pregnancy. On other hand, the study will not request the name of the respondent. All information that will be obtained from the study will be secured and used for academic purpose.

To ensure voluntary participation the study will give the participants chance to withdraw from the study and not being subjected to any negative consequence. Being pregnant during adolescence period make these girls to be vulnerable to exploitations so this study will ensure that adolescent pregnant girls are not exploited in any form either physically or physiologically.

### Limitation of the study

The study will have the following limitations, first its only adolescent mother will participate in this study so the results might not be generalized to adolescent girls who do not have children second the study will be performed in rural areas so those who live in the Dodoma city will not be able to be part of it hence it will affect generatability of the study findings. Third, Adolescent mothers who are not be able to speak Swahili will be excluded from the study, this will lead to miss some of important information. Besides no pregnancy test will be carried out to prove the existence of pregnancy so there is a chance to miss adolescent pregnant mothers below 12 weeks of gestational age. Last, since the study adapted quantitative approach, structured questionnaire will be employed this will give no room for in depth information about the adolescent pregnant mothers on factor that affected their knowledge attitude and health seeking behaviour during their pregnancy.

### Dissemination plan

Findings obtained from this study will be disseminated to the University of Dodoma, the Ministry of health, P0-RALG and will be submitted for publication in peer-reviewed journal.

### How amendments to the study, including termination, will be dealt with

If any changes are made to the study, including the study’s termination, this will be communicated to the journal auditoria, which will submit the changes or provide a reason for the change.

## Data Availability

No datasets were generated or analysed during the current study. All relevant data from this study will be made available upon study completion.

## Authors contribution

Writing – original draft, Conceptualization and Methodology: Eliwedi Mbwambo Writing – review, editing and Supervision: Nyasiro Gibore

## Acknowledge

To my research partners, who put in a lot of work alongside me, I would want to express my sincere gratitude. Their contributions, debates, and ideas greatly enhanced this research. With deep gratitude, I also acknowledge Dr. Nyasiro Gibore for her steadfast advice during this research process.

